# The Psychosocial Benefits of Sport Participation During COVID-19 Are Only Partially Explained by Increased Physical Activity

**DOI:** 10.1101/2022.01.11.22269077

**Authors:** Andrew M. Watson, Kevin Biese, Claudia Reardon, Allison Schwarz, Kristin Haraldsdottir, M. Alison Brooks, David R. Bell, Timothy McGuine

## Abstract

The purpose of this study was to determine whether physical activity (PA) increases were responsible for the improvements in mental health and quality of life (QOL) seen among adolescents who returned to sport during the COVID-19 pandemic.Adolescent athletes were asked to complete a survey in October 2020 regarding demographic information, whether they had returned to sport participation (no [DNP], yes [PLY]), school instruction type (virtual, in-person, hybrid), anxiety, depression, QOL, and PA. Anxiety, depression, QOL and PA were compared between PLY and DNP using least squares means from linear models adjusted for age, gender, and instruction type. Mediation analysis assessed whether the relationship between sport status and anxiety, depression, and QOL was mediated by PA. 171 athletes had returned to play, while 388 had not. PLY athletes had significantly lower anxiety (3.6±0.4 v 8.2±0.6, p<0.001) and depression (4.2±0.4 v 7.3±0.6, p<0.001), and significantly higher QOL (88.1±1.0 v 80.2±1.4, p<0.001) and PA (24.0±0.5 v 16.3±0.7, p<0.001). PA explained a significant, but relatively small portion of the difference in depression (22.1%, p=0.02) and QOL (16.0%, p=0.048) between PLY and DNP athletes, but did not explain the difference in anxiety (6.6%, p=0.20). Increased PA is only responsible for a small portion of the improvements in depression and QOL among athletes who returned to sports and unrelated to improvements in anxiety. This suggests that the majority of the mental health benefits of sport participation for adolescents during the COVID-19 pandemic are independent of, and in addition to, the benefits of increased PA.

## INTRODUCTION

The cancelation of school and sports during the COVID-19 pandemic has been associated with significant decreases in physical activity and worsening mental health and quality of life.[1-5] In a study of over 13,000 adolescent athletes in May 2020, shortly after the nationwide cancelation of school and sports, 37% reported moderate to severe symptoms of anxiety and 40% reported moderate to severe symptoms of depression.[4] When the subset of this group of adolescent athletes from Wisconsin were compared to historical data collected from adolescent Wisconsin athletes prior to the pandemic, it was found that athletes restricted from sports during the COVID-19 lockdown reported significantly worse QOL and dramatically higher symptoms of anxiety and depression, even after adjusting for age, gender, and school instruction method.[6]

Physical activity (PA) has been consistently demonstrated to have significant mental health benefits, and sports participation may also offer psychosocial benefits that are independent of, and in addition to, the benefits of increased PA.[7-9] For example, sport participation is associated with significant psychological and social health benefits, and athletes demonstrate higher QOL and self-esteem than their non-athlete counterparts, as well as greater academic success.[9-11] Recent research has found that survivors of the Severe Acute Respiratory Syndrome outbreak in 2002-2003 demonstrate a significantly increased prevalence of mental illness twenty years later.[12] Given the worsening mental health epidemic among adolescents during COVID-19, this suggests that interventions to reduce the mental health impacts of the current pandemic could have vitally important long-term benefits among adolescents.

Prior research has found that adolescent athletes who returned to participation in organized sports had significantly higher PA and QOL, and significantly lower symptoms of anxiety and depression than athletes who were unable to return to sports.[13] It remains unclear, however, whether the psychosocial benefits of returning to sports that were identified are due to the increased PA or other facets of sport participation such as the restoration of social networks or athletic identity. This can provide valuable information to help inform discussions regarding the re-initiation of organized sports during the COVID-19 pandemic. Therefore, the purpose of this study was to conduct a secondary analysis of previously published data[13] to determine whether increases in PA mediate the psychosocial benefits of returning to sports among adolescent high school athletes. We hypothesized that physical activity differences between athletes who did or did not return to sports in the fall of 2020 would mediate a portion of the increased QOL, anxiety and depression, but not a majority of the difference.

## MATERIALS AND METHODS

This study was approved by the University of Wisconsin Health Sciences Institutional Review Board in September 2020. Wisconsin high school athletes (male and female, grade: 9–12, age: 13-19) were recruited to participate in the study by completing an anonymous online survey in October 2020. Emails were sent to athletic trainers and coaches from a convenience sample of 44 schools to solicit their athletes to participate in the study. The survey included a section to solicit demographic information, as well as measures of PA, mental health and QOL. Demographic responses were obtained regarding the participant’s age, sex, grade, school name and whether the athlete planned to participate in their respective sport if it was offered in the 2020-21 school year.

The remainder of the survey consisted of an assessment of mental health, PA and QOL. The General Anxiety Disorder-7 Item (GAD-7) and Patient Health Questionnaire-9 Item (PHQ-9) surveys were used to evaluate anxiety and depression symptoms. The questionnaires ask participants to rate the frequency of anxiety or depression symptoms experienced in the past two weeks. The GAD-7 scale is a valid, reliable and sensitive measure of anxiety symptoms and is able to differentiate between mild and moderate GAD in adolescents. [14] Scores range from 0-21 with a higher score indicating increased anxiety. In addition to the total score, GAD-7 categorical scores of 0–4, 5-9, 10–14, and 15–21 correspond to no, mild, moderate, and severe anxiety symptoms, respectively. The PHQ-9 is a 9-item screening questionnaire for depression symptoms with scores ranging from 0-27 with a higher score indicating a greater level of depression.[15]

PA was assessed with the Hospital for Special Surgery Pediatric Functional Activity Brief Scale (PFABS). This represents a measure of overall physical activity that has been validated in adolescents[16] and has published normative adolescent data.[17] QOL was measured with Pediatric Quality of Life Inventory 4.0 (PedsQL). The type of instructional delivery method (online only, in person or hybrid [a combination of in person and online]) was determined by reviewing information on each school’s website. Participants were excluded if they did not complete the entire survey, were not in grades 9-12, or indicated they did not plan to play interscholastic sports at their school for reasons other than COVID-19 restrictions. Participants were classified as playing a fall sport (PLY) or as not playing a fall sport (DNP).

Data were initially grouped by sport status (DNP, PLY). To evaluate the association between sport status and anxiety, depression, QOL, and PA, least squares means from separate linear regression models adjusted for age, sex, and school instruction type were used to compare groups. The relationship between PA and anxiety, depression, and QOL was evaluated using similar adjusted regression models. In order to identify the relationship between PA and psychosocial outcomes, PA was included in separate similarly adjusted linear models to predict anxiety, depression, and QOL. Finally, separate mediation analyses were conducted to evaluate the proportion of the difference in anxiety, depression, and QOL between the DNP and PLY groups that was explained by differences in PA (see Figure 1).

**Figure 1.**
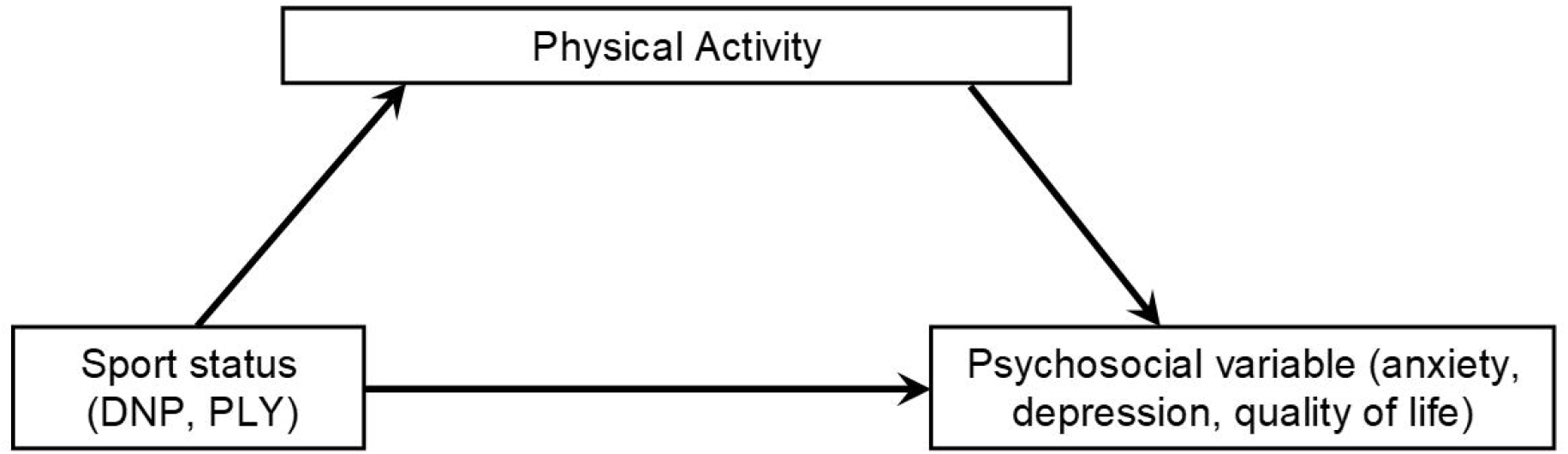
Mediation analysis, demonstrating the independent variable (sport status), the mediator variable (physical activity), and the dependent psychosocial variable (anxiety, depression, or quality of life). This approach evaluates the proportion of the effect of the independent variable on the dependent variable that is explained by the mediator variable. DNP = did not play sports in fall 2020. PLY = did play sports in fall 2020.

Specifically, for each psychosocial outcome variable, two models were developed: 1) a linear model to predict the variable with age, sex, school instruction type and sport status as covariates, and 2) a linear model to predict the variable with PA, age, sex, school instruction type and sport status as covariates. Using the mediate() function in R, the outputs from the 2 models were used to generate 500 quasi-Bayesian Monte Carlo simulations that yield parameter estimates and 95% confidence intervals. Statistical significance in the final mediation analysis was considered *a priori* at p < .05, and all tests were 2-tailed. Data are presented as n (%) for categorical variables and mean(standard deviation) for continuous variables. Statistical analyses were performed in R.[18]

## RESULTS

As previously reported, a total of 559 high school athletes (age = 15.7+1.2 yrs., female = 43.6%, male = 56.4%) completed the survey. Three hundred eighty-eight (69.4%) participants reported they did not play (DNP) an interscholastic sport at their school, while 171 (30.6%) reported they did play (PLY) an interscholastic sport. PLY athletes were found to have higher PA and QOL, and lower depression and anxiety than DNP (see Table 1). PA was significantly and positively related to QOL and significantly, inversely related to anxiety and depression symptoms (see Table 2). Finally, PA was found to mediate a significant portion of the relationship between sport status and depression and QOL, but not anxiety (see Table 3).

**Table 1.** Differences in physical activity, anxiety, depression, and quality of life between adolescent athletes who did or did not return to sport participation in fall 2020.^a^

| Variable | DNP | PLY | p |
| --- | --- | --- | --- |
| Physical Activity (PFABS) | 16.3 ± 0.7 | 24 ± 0.49 | <0.001 |
| Anxiety (GAD-7) | 8.21 ± 0.57 | 3.6 ± 0.4 | <0.001 |
| Depression (PHQ-9) | 7.34 ± 0.61 | 4.18 ± 0.43 | <0.001 |
| Quality of Life (PedsQL) | 80.2 ± 1.4 | 88.1 ± 0.99 | <0.001 |

Data are presented as mean ± SD. <sup>a</sup>Comparisons between groups using least squares means from linear regression models adjusted for age, sex, and school instruction type. DNP = did not return to sport participation; GAD-7 = Generalized Anxiety Disorder 7-Item; PedsQL = Pediatric Quality of Life inventory; PHQ-9 = Patient Health Questionnaire 9-Item; PLY=did return to sport participation.

**Table 2.** Relationship between physical activity and psychosocial outcomes in adolescent Wisconsin athletes in fall 2020.^a^

|  | Estimate | SE | p |
| --- | --- | --- | --- |
| Anxiety (GAD-7) | -0.103 | 0.034 | 0.002 |
| Depression (PHQ-9) | -0.130 | 0.035 | <0.001 |
| Quality of Life (PedsQL) | 0.269 | 0.081 | <0.001 |

<sup>a</sup>Relationships evaluated by linear regression models adjusted for age, gender, and school instruction type. DNP: did not return to sport participation; GAD-7 = Generalized Anxiety Disorder 7-Item; PedsQL = Pediatric Quality of Life inventory; PHQ-9 = Patient Health Questionnaire 9-Item.

**Table 3.** Proportion of the difference in psychosocial outcomes between adolescent athletes who did or did not return to sport participation in fall 2020 that is explained by physical activity.

|  | Proportion Mediated by PA | Lower CI | Upper CI | p |
| --- | --- | --- | --- | --- |
| Anxiety (GAD-7) | 6.6% | -3.4% | 19.5% | 0.196 |
| Depression (PHQ-9) | 22.1% | 3.8% | 74.3% | 0.020 |
| Quality of Life (PedsQL) | 16.0% | 0.29% | 46.6% | 0.048 |

CI = 95% confidence interval; GAD-7 = Generalized Anxiety Disorder 7-Item; PA = physical activity; PedsQL = Pediatric Quality of Life inventory; PHQ-9 = Patient Health Questionnaire 9-Item.

## DISCUSSION

Sport participation has been associated with a number of beneficial physical and mental health outcomes for adolescents, as well as higher academic success.[9, 11] Following the widespread sport and school cancelations in response to the COVID-19 pandemic in the spring of 2020, adolescent athletes demonstrated decreases in PA and QOL, as well as marked increases in anxiety and depression.[4] In fall 2020, athletes who were able to return to sports demonstrated higher PA and QOL that approached historical, pre-pandemic values, as well as significantly better mental health scores compared to those athletes unable to return to sports.[3] Specifically, after adjusting for age, sex, school instruction type and socioeconomic status, athletes who were unable to return to sports were more than 6 times as likely to report moderate to severe anxiety and more than twice as likely to report moderate to severe symptoms of depression.[13] This is consistent with prior research that has found that social connections through sports have an important influence on mental health in student-athletes,[19] and that student-athletes with more social support and connectedness had less dissolution of athletic identity and improved mental health.[20] In this study, we demonstrate that increases in PA among those who returned to sports explained only a small portion of the overall benefits of sports participation on mental health and QOL among adolescent athletes during the COVID-19 pandemic. Specifically, we found that PA explained 22% of the difference in depression and 16% of the difference in QOL between PLY and DNP athletes. In addition, PA explained only 7% of the difference in anxiety, which was not statistically significant. This seems to align with prior research suggesting that increased PA is associated with improvements in a wide range of psychosocial outcomes, but suggests that the majority of the difference in mental health and QOL between DNP and PLY athletes in the current study is attributable to aspects of sport participation beyond just increased levels of PA. It also suggests that the increased anxiety experienced by student-athletes unable to return to sports in the fall of 2020 sports is largely unrelated to the loss of physical activity. While we cannot directly address the underlying cause, this may be primarily attributable to other factors, such as loss of athletic identity, uncertainty regarding the future of their sports career, increased exposure to negative home or peer environments without time in sports, and/or the loss of social connections that sports provide.[19, 20]

Participation in organized sports can offer social connections, interactions with peer networks and role models, as well as a broader sense of purpose and identity for adolescents.[21] During the COVID-19 pandemic, organized sports may offer an even more pronounced influence as a means to combat social isolation and the pervasive sense of uncertainty that surrounds the cancelation of “normal” activities for children and adolescents. Here we demonstrate that the myriad psychosocial benefits of sport participation are not only significant during the COVID-19 pandemic, but that they are only partly attributable to PA. This suggests that while efforts to increase or maintain PA may be helpful in reducing symptoms of depression or improving quality of life through the pandemic, the re-initiation of youth sports can have even greater benefits for QOL and mental health among adolescent athletes.

This study has several limitations. Although we attempted to account for differences between the groups with respect to age, sex, and school instruction type through adjusted models, it is possible that other factors that differ between the DNP and PLY groups are not accounted for could confound our results. We cannot be certain that the differences in PA are entirely explained by participation in sports as athletes in both groups may have sources of PA outside of sports. We did not directly measure differences between groups with respect to factors that could potentially influence psychosocial outcomes, such as the social benefits or athletic identity, and can only speculate about the role that these may play in influencing the differences between groups. It is unknown if the relationships between PA and psychosocial outcomes in adolescent athletes during the COVID-19 pandemic will differ after the pandemic is over. Finally, this analysis represents a group of adolescent athletes from a single state, which may not be generalizable to other populations.

## CONCLUSIONS

In summary, return to participation in sports during the COVID-19 pandemic is associated with higher PA, improved QOL, and reduced symptoms of anxiety and depression in adolescent athletes. We found that although increased PA is associated with improved QOL and reduced anxiety and depression symptoms, it only explains a small portion of the difference in these outcomes between those athletes that did or did not return to sports. This suggests that elements of organized sport participation beyond PA play an important role in helping improve psychosocial outcomes for adolescent athletes during the COVID-19 pandemic. While PA can have psychosocial benefits for adolescents during the pandemic, participation in sports may offer greater benefits than PA alone. This information may help inform stakeholders regarding the re-initiation and/or continuation of organized youth sports during COVID-19.

## Data Availability

Data are available from the study team upon reasonable request.

## ACKNOWLEDGEMENTS

There are no funding sources to report or competing interests to declare for this study. Dr. Watson is supported by grants from the National Center for Advancing Translational Sciences (UL1TR002373; KL2TR002374). We are grateful for the resources and support of the UW Institute for Clinical and Translational Research.

